# “From Preparation to Consumption.” Food Safety Practices among Public Food Handler’s in Enugu Metropolis

**DOI:** 10.1101/2023.10.10.23296821

**Authors:** Chukwukasi Wilson Kassy, Nwadiuto Chidinma Ojielo, Ugenyi Victoria Iloabachie, Casmir Ndubuisi Ochie, Ifeoma Juliet Ogugua, Ibiok Charles Ntat, Onyinye Hope Chime, Chioma Amarachi Onyedinma, Prof Anne Chigedu Ndu, Prof Uzoamaka Susan Arinze-Onyia, Prof Nwabueze Emmanuel Aguwa, Prof Adaeze Theodora Okeke

## Abstract

**Background:** Increased demand for public prepared food with numerous food handlers creates uncertainties in the quality of safe foods and possible food contamination. This study aimed to ascertain the food safety hygiene practices and associated factors among public food handlers in Enugu Metropolis, Nigeria.

**Methodology:** This was a cross-sectional study conducted among 400 public food handlers in Enugu Metropolis, Nigeria. This study took place between January and April 2023. Samples were selected using multistage sampling technique. Data was collected using pretested structured questionnaire and analyzed using proportion, mean and multiple regression. Statistical significance was set at < 0.05.

**Results:** The mean age of respondents were 31.16 ± 8.242 years. About two – thirds, 66.5% of respondents were found to have good knowledge of food hygiene safety practices. The overall food safety hygiene practice mean score was 80.10 ± 10.25 with 70.5% showing good practice. Environmental safety hygiene had good practice of 35.0% and mean score of 24.17 ± 2.29. The factors which statistically significantly predicted overall food safety practices, F (11, 388) = 42.957, P < 0.0001, R^2^ = 0.536 were educational level (β = 0.148, C.I = 0.860 – 3.082), knowledge level (β = 8.594, C.I = 5.635 – 8.979) and safety trainings (β = 0.517, C.I = 4.102 – 5.474).

**Conclusion:** There was good knowledge of food safety hygiene practices with high mean scores except on environmental safety hygiene practices component. Safety training, knowledge level and educational level were the predictors of good practices. Frequent trainings are most needed to prevent or control food contamination and consequent food borne diseases.

## Introduction

Food safety is the state of certainty that food will not cause harm either from “farm to table” or from “preparation to consumption”. [1] Food safety practice is a public health measure to prevent and control food – borne illnesses caused by bacteria, viruses, parasites and chemicals through contamination of food or water. The practices involves keeping oneself and surfaces clean, separate raw and cook foods, cook food thoroughly, store at safe temperature and use of safe water and raw materials. It has become a public health priority due to urbanization and globalization with increased demand for public prepared food, variety of food, complex and longer global food chain. [2-4] The safety practices comprises of personal hygiene practices such as personal cleanliness, safe handling of food, use of personal protective equipment; environmental hygiene practices such as general cleanliness and upkeep of premises, appropriate layout, adequate lightening, ventilation, pest and waste management and Quality control practices such as Hazard Analysis and Critical Control Points (HACCP), labelling, traceability, staff training.[5] The sum total of all the above practices constitute the Food safety system which is the management system, establishment must have in place if they sell food. [6] It is most applicable in the well developed and established food and beverage production industries, large scale and multi – outlets food companies that ensures no weak link in the “farm to table” or “preparation to consumption” concept of food supply is allowed. However, because of the complexities of the quality control HACCP, small scale retail food outlets and street food vendors are limited to food hygiene without standardized quality control measures.

The “farm to table” concept best summarizes food supply chain management (FSCM) that once food is harvested or produced, should be stored, distributed, retailed and consumed.[7] The complexity to maintain food safety has allowed some companies and food outlets to enter the supply chain at some points mostly from market (retail) to consumption, hence the concept “preparation to consumption” as seen among domestic and public food handlers (away from home food consumption).[8] Food preparation involves activities like purchasing, washing, trimming, peeling, grinding, blanching, boiling, cooking, roasting, frying etc. [9] With increasing population, globalization and urbanization, there is consequent increase in public food consumption (the away – from – home food consumption) from sources like fast food, cafeterias and restaurants. The food preparation are done either in indoor kitchen or outdoor kitchen premises where the environmental hygiene practices could be a concern for contaminations. [10] Studies in Nigeria found a mix of poor and good knowledge of food safety hygiene practices with poor regulatory system in the food supply chain among domestic and / or public food handlers. [11-13] However, these findings were mainly on personal hygiene practices without much known of environmental hygiene practices, hence, undermining the weight of the safety hygiene practices for adequate safety control measures.

Unsafe contaminated food causes food borne illness of more than 200 diseases ranging from diarrhea to cancer. An estimated 600 million (1 in 10) and 420,000 people worldwide respectively fall ill and die from unsafe food annually resulting in loss of 33 million healthy life years. Children under 5 years contribute 40% of food borne diseases with 125,000 deaths each year. [2] About 10 to 20% of unsafe contaminated food by handlers results in outbreaks of which food poisoning and diarrheal diseases are the commonest. Diarrheal diseases from unsafe contaminated food causes 550 million illness and 230,000 deaths annually. [2,14]. Estimated $110 Billion is lost annually in productivity and medical expenses from unsafe food globally. [2]. In Nigeria, 200,000 people die annually from unsafe food with $3.6 Billion associated with cost of food borne illness. [11,15]

Ensuring adequate food safety control measures among public food handlers, both the personal and environmental food safety hygiene practices must be encouraged. Looking into these practices strengthen the degree of confidence among food handlers and owners that food will not cause harm. It will further fill in the gap on knowledge of the environmental hygiene practiced by food vendors. This study will help create awareness among food regulatory agency and policy makers on the degree of food safety hygiene practiced and help designed a standardized protocol for food hygiene training and practice for food handlers and vendors. This study aimed to ascertain the food safety hygiene practices and associated factors among public food handlers in Enugu Metropolis, Nigeria.

## Methodology

### Study Area

The study was conducted in Enugu Metropolis, the capital of Enugu State, Nigeria. The metropolis is constituted by three Local Government Areas which are Enugu North, Enugu South and Enugu East and is inhabited primarily by the Igbo ethnic group. [16] According to the 2016 estimated population, Enugu Metropolis had a total population of 820,000 representing 22.2% of the Enugu State population (4,411,100) with projected growth of 2.58%. [17,18] The metropolis is an administrative, educational and commercial city where the main occupation are civil service and trade with small scale business ranging from artisans to public services. Among the small scale business is food supply business either as stationed or mobile food outlets. This help to feed the teaming population of the city growth and help working people manage the time demand of their job.

### Study design

The study design is a cross – sectional descriptive study.

### Study population

This comprised of all public food handlers or vendors in Enugu metropolis who have been in the services of food supply for 6 months or more will be selected while those who are less than 6 months in food supply services, are seriously ill or absent from work will not be selected.

### Sample size determination

The minimum sample size was calculated using a sample size determination for a single proportion. [19] The proportion of good food safety practices among food handlers in Owerri, Imo State, Nigeria was 37% [20] and the margin of error was set at 5%. The calculated minimum sample was 393.8 after adjusting for non – response rate. This was approximated to 400 food handlers / vendors to be studied.

### Sample size selection

The total sample frame of public food vendors (hotels, hospital and school cafeterias, restaurants, food kiosks, road side food sellers and food hawkers) in Enugu metropolis were gotten from the trade unions and local governments. A proportionate sampling followed by simple random sampling was used to select the required samples size for the study.

## Data collection

The study started on the 22^nd^ of January,2023 and end of recruitment of this study was on the 29^th^ of April 2023. The duration of this study was 4 months. A structured questionnaire pretested in the local government not selected was used. The questionnaire was adapted from the literature. (References 1 – 5). It contains the socio – demographic, knowledge and practice sections. The data was collected using the aid of research assistance who were trained on the objectives and ethics of the study. They were trained for two days, 2 hours per day.

## Data analysis

Data were manually clean, entered and analyzed using IBM Statistical Produce and Service Solution (SPSS) version 25. The Knowledge questions has two responses, the right response was Yes while the wrong response was No. the right is coded as 1 while the wrong responses as 0. The total score is 14, those with score of 50% and above was noted as good knowledge while those with less than 50% was were noted as poor knowledge. The practice questions has responses in a Likert scale of never, rarely, sometime, often and always and were coded respectively as 1, 2, 3, 4 and 5. There were a total of 23 questions with total score of 115. Scores were summarized using mean and standard deviation. Those with scores above 60% were regarded as having good practice while those with scores of 60% and less as poor practice. Categorical variables were summarized using frequency table and proportions. Determinants of good practice were analyzed using multiple regression analysis.

### Ethical Clearance

This was obtained from the Health Research Ethics Committee of University of Nigeria Teaching Hospital, Ituku / Ozalla, Enugu State, Nigeria. Informed consent were obtained from the food handlers.

## Results

The mean age of respondents were 31.16 ± 8.242 years with age ranging from 18 to 60 years. More than half, 59.3% of respondents were females and about 41.5% had secondary education as the highest educational level. Majority, 64.5% of the respondents were fully employed and more than one – fifth, 84.5% had stayed between one to ten years in their job.

About two – thirds, 66.5% of respondents were found to have good knowledge of food hygiene safety practices. More than three – quarter, 79.5 and 79% respectively noted that food equipment should be washed before or immediately after use, and hands frequently washed with soap and water. While less than one – third, 29.0 noted that contaminated food stuffs cannot be detected using sense organs.

The overall food safety hygiene practice mean score among public food handlers was 80.10 ± 10.25 with 70.5% showing good practice. Among the subscales, public food handlers practicing workplace safety hygiene had good practice of 66.5% and mean score of 28.52 ± 3.73, personal safety hygiene had good practice of 62.5% and mean score of 27.40 ± 5.70 and environmental safety hygiene had good practice of 35.0% and mean scores of 24.17 ± 2.29.

The factors which statistically significantly predicted overall food safety practices, F (11, 388) = 42.957, P < 0.0001, R^2^ = 0.536 were educational level (β = 0.148, C.I = 0.860 – 3.082), knowledge level (β = 8.594, C.I = 5.635 – 8.979) and safety trainings (β = 0.517, C.I = 4.102 – 5.474). For the subscales, environmental safety health practices F (11, 388) = 8.973, P < 0.0001, R^2^ = 0.203 was safety trainings (β = 0.397, C.I = 0.619 – 1.027), for personal hygiene safety practices, F (11, 388) = 37.935, P < 0.0001, R^2^ = 0.518 were knowledge level (β = 0.322, C.I = 2.922 – 4.844) and safety trainings (β = 0.534, C.I = 2.356 – 3.145), for workplace safety health practices, F (11, 388) = 45.919, P < 0.0001, R^2^ = 0.360 were educational level (β = 0.256, C.I = 0.849 – 1.644), knowledge level (β = 0.424, C.I = 2.747 – 3.942) and safety trainings (β = 0.360, C.I = 0.969 – 1.459).

## Discussion

The complexities in carrying out quality control food hygiene has limited small scale public food handlers and vendors to personal and environmental hygiene practices. This study seeks to maximize the impact of these available practices and ensure food safety to the teeming population at all times. The study found about two – third of the respondents have good knowledge of food safety practices. Most of these were driven by knowledge on best food preparation and cooking practices otherwise knowledge on storage of cooked foods in refrigerator were poor. These findings agreed with studies in Nigeria, South Africa, Ethiopia and Brazil. [14, 20-23] These could be due to the increased awareness and health education campaigns on hand hygiene, personal cleanliness and environmental sanitation globally including infection, prevention and control in health and health related activities.

Consequently, the study found higher mean scores on the overall and subscales food safety hygiene practices. With about two – thirds of the practices on the overall food safety hygiene, personal safety hygiene and workplace safety hygiene subscales found to be good practices, however, about one – third of the practices in the environmental subscale were good. The poor practices found in the environmental subscale or component will continue to be a source of contamination in the food processes chain, hence, leading to unsafe food and consequent food borne diseases. It further shows that the weight of a component of the food safety hygiene practices may not be enough to tilt the scores to the left but practically could constitute to unsafe food. These findings disagreed with studies in Nigeria, Northwest Ethiopia but agreed with study in Brazil with very high proportion of good safety practices. [14, 20, 22, 23] Despite the difference and or similarity, the current findings methodologically, was different from previous studies as the contributions of the component subscales were taken into consideration. Also the responses were graded, that showed the weight of the safety practices. The public health importance of the findings is that safety practices scores of above 60% means practices that are “often and always” done to which minimizes the risk of food contamination, otherwise scores below such will maximize the risk of food contamination as safety practices are “sometimes, rarely and never” done. Hence, environmental safety hygiene practices are potential risk for contamination in the study population. More public health intervention measures through food safety trainings should be enforced targeting more of environmental practices, as this was “sometimes, rarely and never” done in this study.

The study further found that safety training, knowledge level and educational level were the predictors of good practices in the overall and subscales food safety hygiene practices. Safety training was the commonest predictor of food safety hygiene practices as it was consistent in both the overall scale and the subscales. This was found to agree with study in Ethiopia [24]. This is because with frequent trainings on food safety hygiene practices, there will be improved awareness and knowledge of the various safety practices including the degree of such practices. Hence, the need for frequent safety training in food safety management system from government to individual levels.

## Conclusion

The study found that there is good knowledge of food safety hygiene practices, high mean scores in both the overall and subscales with good safety hygiene practices except on environmental safety hygiene practices component. Safety training, knowledge level and educational level were the predictors of good practices in the overall and subscales food safety hygiene practices.

### Recommendation

Frequent trainings done often or always are needed as public health measures to maintain the level of food safety hygiene practices that will prevent or control food contamination and consequent food borne diseases.

### Limitations

The study is limited to one site and therefore the results are not generalizable

### Funding

Authors received no funding

## Data Availability

All relevant data are within the manuscript and its supporting information files

---

**Table 1:**
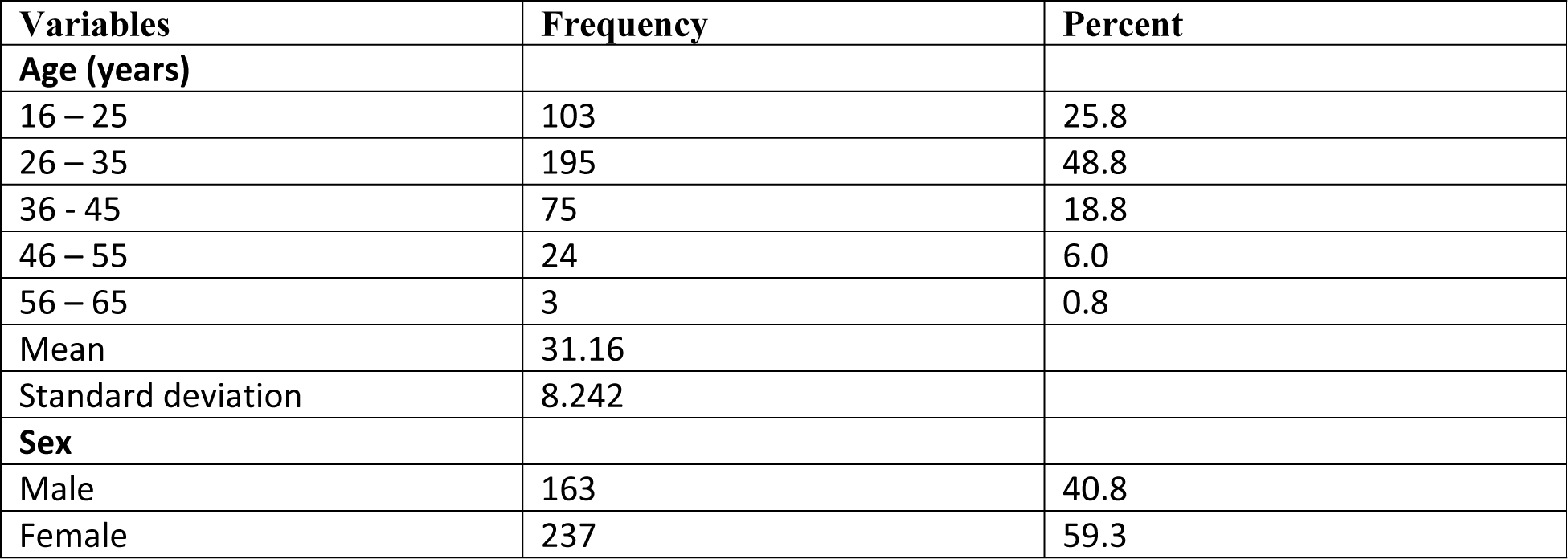
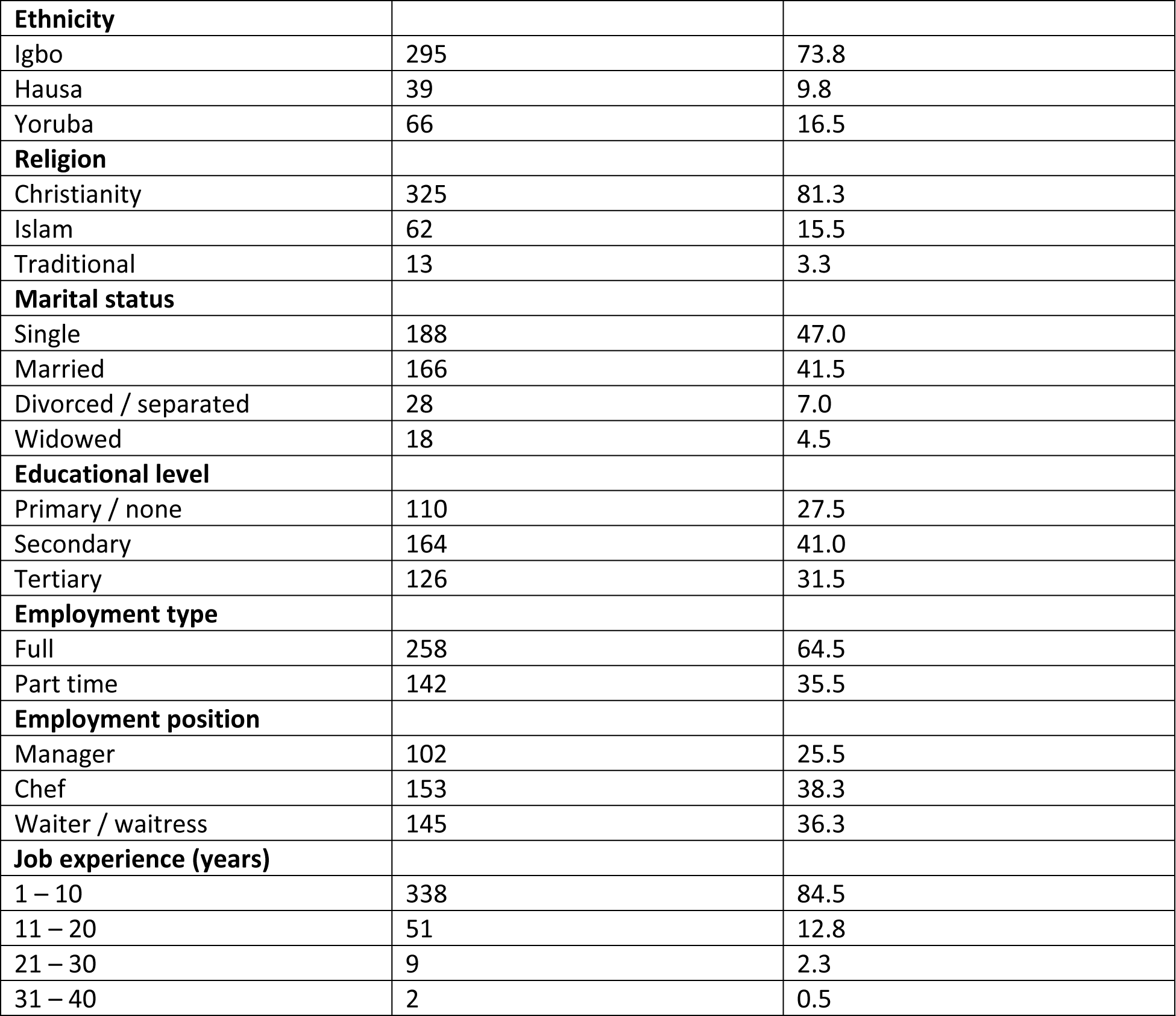
Socio – demographic and Occupational characteristics of respondents.

**Table 2:** Knowledge of Food safety practices among respondents.

| **Variables** | **Frequency** | **Percent** |
| --- | --- | --- |
| Uncooked meat should be stored in lower part of refrigerator | 253 | 63.3 |
| Chilled foods is best stored at refrigerating temperature (5°C or below | 185 | 46.3 |
| Frozen foods is best stored at freezing temperature | 202 | 50.5 |
| Temperature of 5°C or below slows or stop microbes from growing. | 213 | 53.3 |
| Maximum duration for chilled and refrigerated food is 3 – 4 days. | 167 | 41.8 |
| Microbes are killed at boiling or internal cooking temperature (70°C and above) | 309 | 77.3 |
| Temperature at which microbes grows rapidly is danger zone or risk zone (5°C - 60°C) | 316 | 79.0 |
| Contacts between cooked and uncooked foods causes cross – contamination. | 265 | 66.3 |
| Contamination of food stuffs cannot be detected using sense organs (Eyes, nose and tongue). | 116 | 29.0 |
| Wearing gloves, masks and headscarf will reduced contamination of food | 309 | 77.3 |
| Washing hand frequently with soap and water prevents food contamination. | 316 | 79.0 |
| Food equipment should be washed before use or immediately after use. | 318 | 79.5 |
| Contact surface cleaning should be done frequently | 278 | 69.5 |
| Commonest food borne disease is diarrhea. | 207 | 51.8 |
| **Knowledge level** |  |  |
| Poor knowledge | 135 | 33.8 |
| Good knowledge | 265 | 66.3 |

**Table 3:**
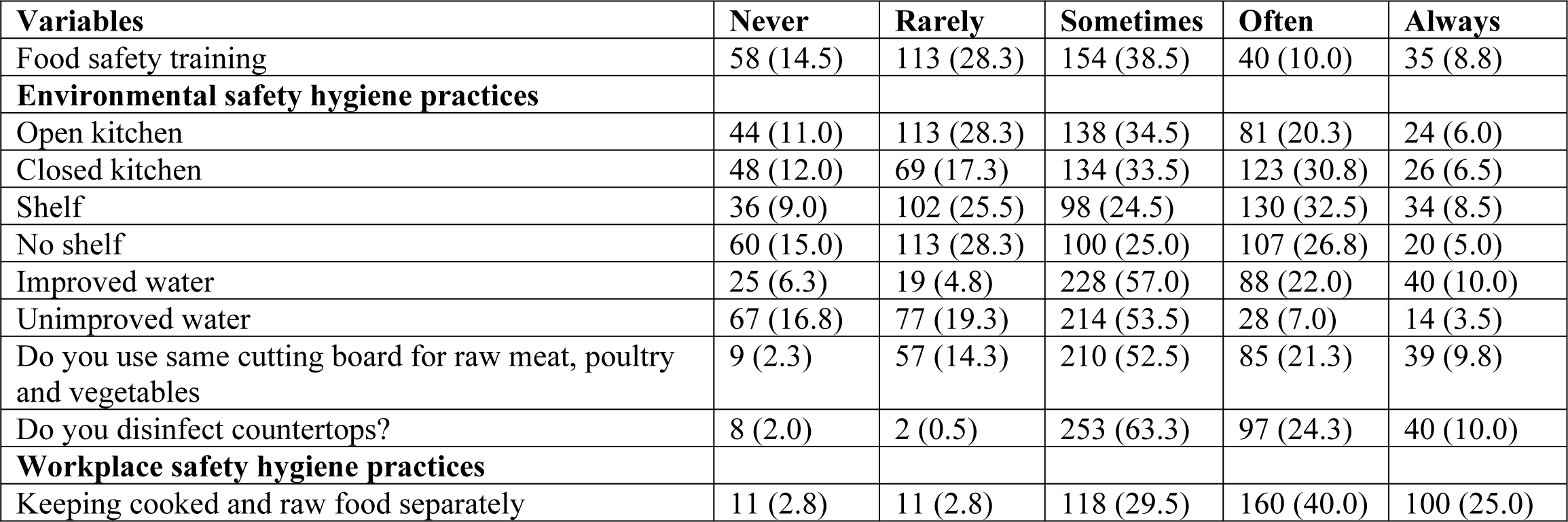
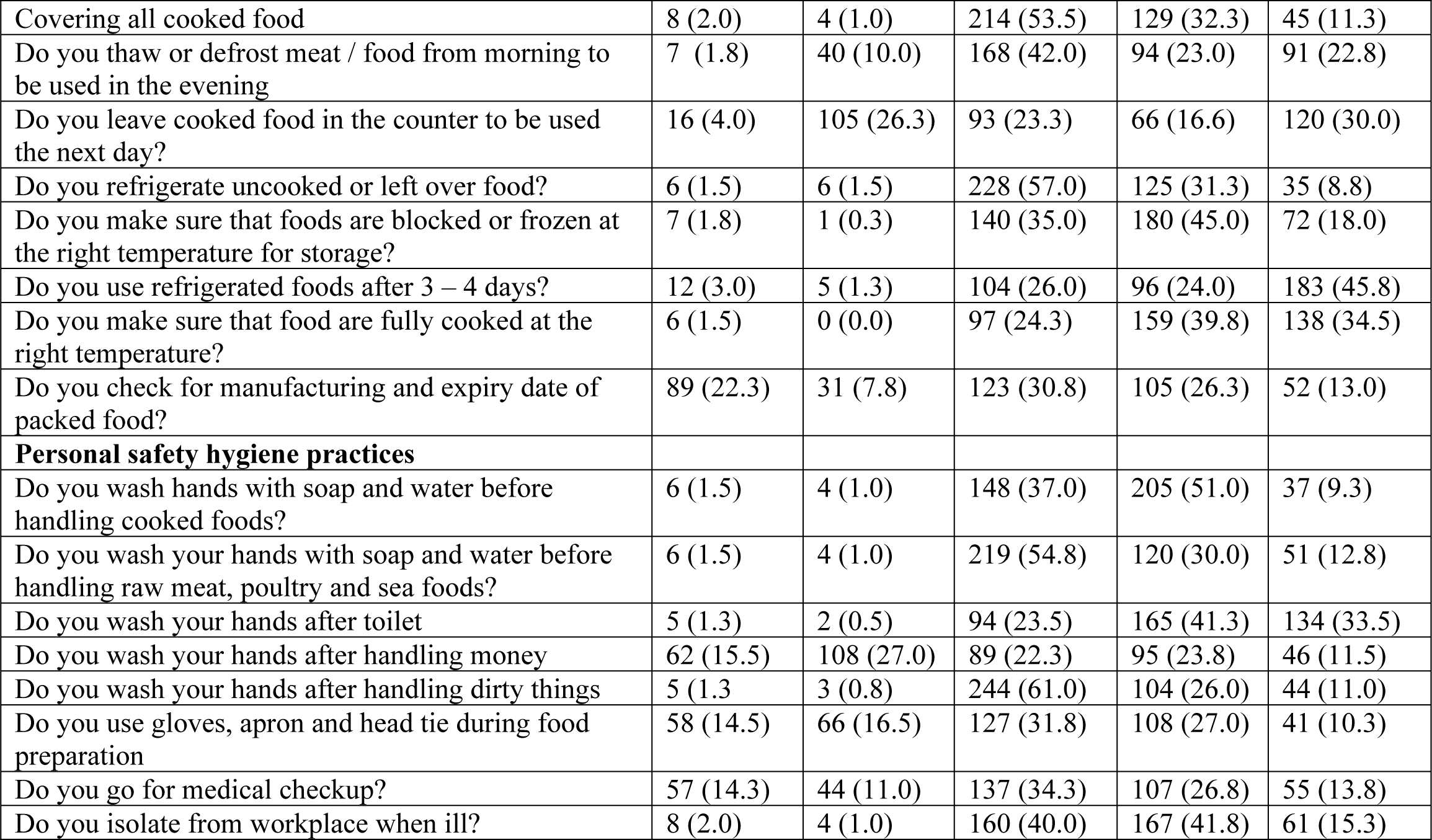
Food hygiene practices.

| **Variables** | **Never** | **Rarely** | **Sometimes** | **Often** | **Always** |
| --- | --- | --- | --- | --- | --- |
| Food safety training | 58 (14.5) | 113 (28.3) | 154 (38.5) | 40 (10.0) | 35 (8.8) |
| **Environmental safety hygiene practices** |  |  |  |  |  |
| Open kitchen | 44 (11.0) | 113 (28.3) | 138 (34.5) | 81 (20.3) | 24 (6.0) |
| Closed kitchen | 48 (12.0) | 69 (17.3) | 134 (33.5) | 123 (30.8) | 26 (6.5) |
| Shelf | 36 (9.0) | 102 (25.5) | 98 (24.5) | 130 (32.5) | 34 (8.5) |
| No shelf | 60 (15.0) | 113 (28.3) | 100 (25.0) | 107 (26.8) | 20 (5.0) |
| Improved water | 25 (6.3) | 19 (4.8) | 228 (57.0) | 88 (22.0) | 40 (10.0) |
| Unimproved water | 67 (16.8) | 77 (19.3) | 214 (53.5) | 28 (7.0) | 14 (3.5) |
| Do you use same cutting board for raw meat, poultry and vegetables | 9 (2.3) | 57 (14.3) | 210 (52.5) | 85 (21.3) | 39 (9.8) |
| Do you disinfect countertops? | 8 (2.0) | 2 (0.5) | 253 (63.3) | 97 (24.3) | 40 (10.0) |
| **Workplace safety hygiene practices** |  |  |  |  |  |
| Keeping cooked and raw food separately | 11 (2.8) | 11 (2.8) | 118 (29.5) | 160 (40.0) | 100 (25.0) |
| Covering all cooked food | 8 (2.0) | 4 (1.0) | 214 (53.5) | 129 (32.3) | 45 (11.3) |
| Do you thaw or defrost meat / food from morning to be used in the evening | 7 (1.8) | 40 (10.0) | 168 (42.0) | 94 (23.0) | 91 (22.8) |
| Do you leave cooked food in the counter to be used the next day? | 16 (4.0) | 105 (26.3) | 93 (23.3) | 66 (16.6) | 120 (30.0) |
| Do you refrigerate uncooked or left over food? | 6 (1.5) | 6 (1.5) | 228 (57.0) | 125 (31.3) | 35 (8.8) |
| Do you make sure that foods are blocked or frozen at the right temperature for storage? | 7 (1.8) | 1 (0.3) | 140 (35.0) | 180 (45.0) | 72 (18.0) |
| Do you use refrigerated foods after 3 – 4 days? | 12 (3.0) | 5 (1.3) | 104 (26.0) | 96 (24.0) | 183 (45.8) |
| Do you make sure that food are fully cooked at the right temperature? | 6 (1.5) | 0 (0.0) | 97 (24.3) | 159 (39.8) | 138 (34.5) |
| Do you check for manufacturing and expiry date of packed food? | 89 (22.3) | 31 (7.8) | 123 (30.8) | 105 (26.3) | 52 (13.0) |
| **Personal safety hygiene practices** |  |  |  |  |  |
| Do you wash hands with soap and water before handling cooked foods? | 6 (1.5) | 4 (1.0) | 148 (37.0) | 205 (51.0) | 37 (9.3) |
| Do you wash your hands with soap and water before handling raw meat, poultry and sea foods? | 6 (1.5) | 4 (1.0) | 219 (54.8) | 120 (30.0) | 51 (12.8) |
| Do you wash your hands after toilet | 5 (1.3) | 2 (0.5) | 94 (23.5) | 165 (41.3) | 134 (33.5) |
| Do you wash your hands after handling money | 62 (15.5) | 108 (27.0) | 89 (22.3) | 95 (23.8) | 46 (11.5) |
| Do you wash your hands after handling dirty things | 5 (1.3) | 3 (0.8) | 244 (61.0) | 104 (26.0) | 44 (11.0) |
| Do you use gloves, apron and head tie during food preparation | 58 (14.5) | 66 (16.5) | 127 (31.8) | 108 (27.0) | 41 (10.3) |
| Do you go for medical checkup? | 57 (14.3) | 44 (11.0) | 137 (34.3) | 107 (26.8) | 55 (13.8) |
| Do you isolate from workplace when ill? | 8 (2.0) | 4 (1.0) | 160 (40.0) | 167 (41.8) | 61 (15.3) |

**Table 4:** Food safety hygiene practices among public food handlers in Enugu Metropolis.

| Variables (cronbach alpha) | Practice scores |  |  | Poor practice |  | Good practice |  |
| --- | --- | --- | --- | --- | --- | --- | --- |
|  | Mean | Standard deviation | Min - max | Frequency | Percent | Frequency | Percent |
| Overall Food safety hygiene practice (0.799) | 80.10 | 10.25 | 37 – 103 | 118 | 29.5 | 282 | 70.5 |
| Environmental safety hygiene practices (0.613) | 24.17 | 2.29 | 8 – 30 | 260 | 65.0 | 140 | 35.0 |
| Personal safety hygiene practices (0.877) | 27.40 | 5.70 | 8 – 40 | 150 | 37.5 | 250 | 62.5 |
| Workplace safety hygiene practices (0.414) | 28.52 | 3.73 | 20 – 39 | 134 | 33.5 | 266 | 66.5 |

\*good practice are scores above 60% (those who often and always practices food safety hygiene)

**Table 6a.** Determinants of Overall food and environmental safety hygiene practices and among public food vendors in Enugu Metropolis.

| **Variables** | **B** | **Std. Error** | **$\beta$** | **T – test** | **P – value** | **Lower C.I** | **Upper C.I** |
| --- | --- | --- | --- | --- | --- | --- | --- |
|  | **Overall food safety hygiene practices** |  |  |  |  |  |  |
| Constant | 47.375 | 3.631 |  | 13.048 | <0.0001 | 40.237 | 54.514 |
| Age | 0.059 | 0.071 | 0.047 | 0.828 | 0.408 | -0.080 | 0.198 |
| Sex | -0.912 | 0.733 | -0.044 | -1.245 | 0.214 | -2.353 | 0.529 |
| Ethnicity | 0.364 | 0.553 | 0.027 | 0.660 | 0.510 | -0.722 | 1.451 |
| Religion | -0.214 | 0.869 | -0.010 | -0.246 | 0.806 | -1.922 | 1.495 |
| Marital status | 0.231 | 0.636 | 0.018 | 0.363 | 0.717 | -1.020 | 1.481 |
| Educational level | 1.971 | 0.565 | 0.148 | 3.487 | 0.001* | 0.860 | 3.082 |
| Employment type | 1.227 | 0.908 | 0.058 | 1.351 | 0.178 | -0.559 | 3.013 |
| Employment position | 0.097 | 0.662 | 0.007 | 0.146 | 0.884 | -1.205 | 1.398 |
| Job experience | 0.106 | 0.090 | 0.058 | 1.180 | 0.239 | -0.071 | 0.283 |
| Knowledge level | 7.307 | 0.850 | 0.338 | 8.594 | <0.0001* | 5.635 | 8.979 |
| Safety training | 4.788 | 0.349 | 0.517 | 13.722 | <0.0001* | 4.102 | 5.474 |
| R = 741 R<sup>2</sup> = 0.536 F(11, 388) = 42.957, P < 0.0001 |  |  |  |  |  |  |  |
| **Environmental safety hygiene practices** |  |  |  |  |  |  |  |
| Constant | 19.664 | 1.080 |  | 18.199 | <0.0001 | 17.539 | 21.788 |
| Age | -0.036 | 0.021 | -0.128 | -1.696 | 0.091 | -0.077 | 0.006 |
| Sex | 0.166 | 0.218 | 0.036 | 0.760 | 0.448 | -0.263 | 0.595 |
| Ethnicity | 0.296 | 0.164 | 0.098 | 1.803 | 0.072 | -0.027 | 0.620 |
| Religion | 0.020 | 0.259 | 0.004 | 0.076 | 0.940 | -0.489 | 0.528 |
| Marital status | 0.294 | 0.189 | 0.101 | 1.552 | 0.121 | -0.078 | 0.666 |
| Educational level | 0.322 | 0.168 | 0.108 | 1.912 | 0.057 | -0.009 | 0.652 |
| Employment type | 0.428 | 0.270 | 0.090 | 1.582 | 0.114 | -0.104 | 0.959 |
| Employment position | 0.248 | 0.197 | 0.084 | 1.260 | 0.208 | -0.139 | 0.635 |
| Job experience | 0.043 | 0.027 | 0.105 | 1.600 | 0.110 | -0.010 | 0.096 |
| Knowledge level | 0.080 | 0.253 | 0.016 | 0.315 | 0.753 | -0.418 | 0.577 |
| Safety training | 0.823 | 0.104 | 0.397 | 7.928 | <0.0001* | 0.619 | 1.027 |
| R = 450 R<sup>2</sup> = 0.203 F(11, 388) = 8.973, P < 0.0001 |  |  |  |  |  |  |  |

**Table 6b.** Determinants of personal hygiene and workplace safety health practices and among public food vendors in Enugu Metropolis.

| Variables | B | Std. Error | β | T – test | P – value | Lower C.I | Upper C.I |
| --- | --- | --- | --- | --- | --- | --- | --- |
| **Personal hygiene safety hygiene practices** |  |  |  |  |  |  |  |
| Constant | 11.588 | 2.087 |  | 5.551 | <0.0001 | 7.484 | 15.692 |
| Age | 0.052 | 0.041 | 0.076 | 1.287 | 0.199 | -0.028 | 0.132 |
| Sex | -0.791 | 0.421 | -0.068 | -1.877 | 0.061 | -1.619 | 0.037 |
| Ethnicity | 0.379 | 0.318 | 0.050 | 1.194 | 0.233 | -0.245 | 1.004 |
| Religion | -0.210 | 0.500 | -0.018 | -0.419 | 0.675 | -1.192 | 0.773 |
| Marital status | -0.086 | 0.366 | -0.012 | -0.235 | 0.814 | -0.805 | 0.633 |
| Educational level | 0.403 | 0.325 | 0.054 | 1.239 | 0.216 | -0.236 | 1.042 |
| Employment type | 0.852 | 0.522 | 0.072 | 1.631 | 0.104 | -0.175 | 1.878 |
| Employment position | -0.388 | 0.380 | -0.053 | -1.019 | 0.309 | -1.136 | 0.360 |
| Job experience | 0.037 | 0.052 | 0.037 | 0.718 | 0.473 | -0.065 | 0.139 |
| Knowledge level | 3.883 | 0.489 | 0.322 | 7.943 | <0.0001* | 2.922 | 4.844 |
| Safety training | 2.750 | 0.201 | 0.534 | 13.711 | <0.0001* | 2.356 | 3.145 |
| R = 720 R<sup>2</sup> = 0.518 F(11, 388) = 37.935, P < 0.0001 |  |  |  |  |  |  |  |
|  | **Workplace safety hygiene practices** |  |  |  |  |  |  |
| Constant | 16.124 | 1.298 |  | 12.419 | <0.0001 | 13.571 | 18.676 |
| Age | 0.042 | 0.025 | 0.093 | 1.658 | 0.098 | -0.008 | 0.092 |
| Sex | -0.287 | 0.262 | -0.038 | -1.095 | 0.274 | -0.802 | 0.228 |
| Ethnicity | -0.311 | 0.198 | -0.063 | -1.575 | 0.116 | -0.700 | 0.077 |
| Religion | -0.024 | 0.311 | -0.003 | -0.076 | 0.939 | -0.635 | 0.587 |
| Marital status | 0.023 | 0.227 | 0.005 | 0.100 | 0.920 | -0.424 | 0.470 |
| Educational level | 1.246 | 0.202 | 0.256 | 6.167 | <0.0001* | 0.849 | 1.644 |
| Employment type | -0.053 | 0.325 | -0.007 | -0.162 | 0.872 | -0.691 | 0.586 |
| Employment position | 0.236 | 0.237 | 0.049 | 0.998 | 0.319 | -0.229 | 0.702 |
| Job experience | 0.026 | 0.032 | 0.040 | 0.815 | 0.416 | -0.037 | 0.090 |
| Knowledge level | 3.344 | 0.304 | 0.424 | 10.999 | <0.0001* | 2.747 | 3.942 |
| Safety training | 1.214 | 0.125 | 0.360 | 9.730 | <0.0001* | 0.969 | 1.459 |
| R = 752 R<sup>2</sup> = 0.566 F(11, 388) = 45.919, P < 0.0001 |  |  |  |  |  |  |  |

